# Screening for Cardiac Disease with Genetic risk scoring, Advanced ECG, Echocardiography, Protein Biomarkers and Metabolomics

**DOI:** 10.1101/2021.06.14.21258923

**Authors:** Patrick A. Gladding, Clementina Dugo, Yvonne Wynne, Heather Semple, Kevin Smith, Peter Larsen, Phillip Shepherd, Erica Zarate, Silas Villas-Boas, Todd T. Schlegel

## Abstract

**Introduction:** Screening patients for cardiovascular disease has not been widely advocated due to cost implications and is reserved for high risk or symptomatic patients. We undertook an exploratory study to evaluate the promising low-cost methods for screening, including genetic risk scoring (GRS), advanced ECG (A-ECG), echocardiography and metabolomics.

**Methods:** 78 patients underwent advanced 5-min ECG and echocardiography, including global longitudinal strain (GLS), and echocardiographic calcium scoring (eCS). A GRS of 27 SNPs (GRS27) related to coronary disease and 3 SNPs for atrial fibrillation was used, as well as hs-troponin (Abbott, Singulex, Roche), NTproBNP (Roche) testing and targeted plasma metabolomics using GC-MS. Results were correlated with the presence of coronary artery disease (CAD) (CT coronary angiography (CTCA)), measures of left ventricular hypertrophy (LVH) (echocardiography and CTCA), and LV systolic dysfunction (LVSD) (echocardiography).

**Results:** LV dysfunction was accurately identified by using either A-ECG (AUC 0.97, 0.89 to 0.99) or NTproBNP. eCS demonstrated accurate discrimination of CAD (AUC 0.84, 95% CI 0.72 to 0.92, p < 0.0001. Troponin I (Abbott/Singulex) had the highest sensitivity and accuracy for the detection of LVH measured by either CT or echocardiography (AUC 0.85, 95% CI 0.73 to 0.92), however specificity was reduced by the presence of LV systolic dysfunction. Metabolomics and A-ECG identified underlying abnormal mechanisms related to both LVH (glycine metabolism) and LV dysfunction, (Citric Acid cycle). Metabolomics provided incidental utility by identifying metformin adherence and nutritional biomarkers.

**Conclusion:** A multi-omic approach to screening can be achieved at relatively low cost, and high accuracy, but will need to be evaluated in larger populations to prove its utility.

## Introduction

In recent decades the pressure on healthcare systems from increasing patient referral volumes, an aging population and excessive use of investigations has resulted in prolonged waiting lists and reduced cost efficiencies. Portable ultrasound, genomics, proteomics, metabolomics and information technology have been touted as being solutions to such problems but have yet to be integrated or evaluated in clinical processes. Recent reports have detailed the integration of high volume ‘omic datasets in otherwise well patients (1-4) however this approach has not been used on hospital triaging or symptomatic patients.

Whilst a small proportion of cardiovascular disease is hereditary e.g. familial cardiomyopathy, hypertrophic cardiomyopathy and coronary artery disease, a significant burden of disease results from lifestyle and dietary issues, such as inactivity and highly processed foods. Genetic or genomic testing therefore identifies only part of individual risk often on the basis of lifetime probability of disease. Whereas genomics reflects inherited disease potential from birth, metabolomics provides an instantaneous global assessment of metabolic pathways, and the endproducts of protein enzymatic metabolism. Metabolomics captures not only the effects of lifestyle and diet but also the environmental exposome, including microbiome and its interaction with the human host. Whilst it has been speculated that integrating genomics with clinical data, traditional protein biomarkers and metabolomics might yield better predictions of disease, there are few studies where this has been done (5).

We undertook an exploratory pilot study to evaluate screening in a group of low to intermediate risk patients referred for CT coronary angiography. Included in this were a group of healthy volunteers. Principal goals were to evaluate the utility of a novel echocardiographic calcium score and global longitudinal strain in the prediction of coronary artery disease (6-8), and advanced ECG (A-ECG) algorithms in the detection of left ventricular systolic dysfunction and left ventricular hypertrophy (9-11).

## Methods

### Patients and Clinical factors

The myHealth e-Body study comprised of 200 patients undergoing CT coronary angiogram for low-intermediate risk chest pain. This interim analysis reports on 78 patients from this study, including healthy volunteers who did not undergo CT. Institutional ethics approval was attained prior to the commencement of the study (Health and Disability Ethics approval #15/NTB/34/AM03). All subjects gave written informed consent. Clinical characteristics included demographics, past medical history, 5-min 12-lead ECG for A-ECG measures, a coronary artery and atrial fibrillation genetic risk score, metabolomics, echocardiography, CT coronary angiography and troponin assays. Healthy volunteers did not undergo CT coronary angiography. Clinical factors were extracted from an e-referral system for ordering CT coronary angiography, and from electronic clinical records. As far as possible the collection of data in this study occurred as part of clinical care, and minimally deviated from standard process.

### Biomarkers and Metabolomics

Blood was collected using EDTA tubes. After centrifugation at 3,000 g, for 5-mins plasma was stored at -80°C before being shipped on dry ice to core lab facilities for testing. The 4^th^ generation hs-troponin assays used included Singulex Erenna® SMC troponin I (Limit of detection 0.04ng/L, inter/intraassay coefficient of variation 6% at cTnI 8.3 ng/L), Abbott ArchitectSTAT® troponin I (Limit of detection 1.2 ng/L, interassay coefficient of variation <10% at 4.7 ng/L), and Roche Elecsys® troponin T (Limit of detection 5 ng/L, interassay coefficient of variation 10% at 13 ng/L). NTproBNP was measured using a Roche Elecsys® assay on a Cobas 6000 analyzer (Measuring Range 5 - 35,000 ng/L, intermediate precision 2.9 - 6.1%). More detail regarding the patient casemix and troponin assays used in this project has been discussed elsewhere (12).

### Genotyping

DNA was extracted from Buffy coat using DNA mini extraction kits (Qiagen, Netherlands) and genotyped for the SNPs used in a 27 SNP genetic risk score (GRS27) for coronary artery disease by Mega et al (13) and atrial fibrillation (AF GRS) by Lubitz et al (14), using Sequenom MALDI-TOF mass spectrometry iPLEX technology. The AF GRS was based on three SNPs rs17570669, rs2200733 and rs3853445 (14, 15), of which rs2200733 has been associated with a risk of ischaemic stroke (16-22). CAD GRS27 scores were compared with scores in an acute coronary syndrome cohort (23).

### Electrocardiography

A-ECG analyzed parameters included those derived from signal averaging of all adequately cross-correlated QRS and T complexes by using software originally assembled at NASA (9, 24) to generate results for: (1) several spatial (derived vectorcardiographic or 3-dimensional) ECG parameters, including the spatial mean and peaks QRS-T angles, the spatial ventricular gradient, and various spatial waveform azimuths, elevations, and time-voltages (9), all derived by using the Frank-lead reconstruction technique of Kors *et al* (25); (2) parameters of QRS and T-waveform complexity derived by singular value decomposition (SVD), for example the principal component analysis (PCA) ratio (24) and the dipolar voltage equivalents (9, 26) of the QRS and T waveforms; and (3) the most applicable parameters from the conventional scalar 12-lead ECG. Data from the 5-min ECGs were also processed for multiple measures of both heart rate and QT interval variability (27). All A-ECG parameters studied and their related detailed methods have been described in previous publications (9, 24, 25, 28). Logistic regression and linear discriminant analysis (LDA), a form of machine learning, was used to compare patterns of A-ECG parameters. Serial A-ECG patterns were visualized using a 3D canonical LDA plot, as described elsewhere (29).

### Cardiac Imaging

Echocardiography was performed by a single Level III trained cardiologist, using a Philips CX50 to obtain standard measures such as left ventricular ejection fraction (LVEF) and septal and wall thicknesses. Left ventricular systolic dysfunction (LVSD) was considered present when LVEF<50%. Left ventricular global longitudinal strain was also measured using QLab, and an echocardiographic calcium score was applied using the method described by Corciu et al (6). CTCA was acquired using a 320-slice Aquillon ONE CT scanner (Toshiba Medical Systems). Obstructive coronary artery disease was defined as the presence of coronary stenosis of diameter loss >50%.

### Metabolomics

Samples were received as plasma and stored at -80°C until thawing, extraction and methanol derivatisation, as described previously (30). Gas Chromatography-Mass Spectrometry (GC-MS) was used for identification and semi-quantitation of amino acids (except arginine), organic acids, and fatty acids. GC-MS instrument parameters were based on Smart et al (30), using an Agilent 7890A gas chromatograph coupled to an 5975C inert mass spectrometer with a split/splitless inlet. Data analysis was semi-automated and analysed using Automated Mass Spectral Deconvolution and identification software (AMDIS) against an in-house library of 165 methyl chloroformate derivatised compounds. Compounds that are not included in this library were manually identified using the National Institute of Standards and Technology (NIST) library. Metabolomic data was expressed as relative abundance in reference to an internal standard D4 Alanine. Absolute quantitation was therefore not used.

### Statistics and Pathway analysis

Univariate analysis was performed using the student T-test for continuous variables, and Spearman coefficient for correlations. Receiver-operating characteristic curve (ROC) analysis was used to assess performance of troponin assays at predicting echocardiographic markers by c-statistic and identifying the optimal cutpoint (maximal sensitivity and specificity). A Pearson correlation matrix was generated and visualised using an interactome map of r value relationships. An interactive network was generated to compare the metadata of patients using a Javascript D3 Force layout. The size of the nodes was based on the strength of the edges between nodes which was proportional to the absolute correlation between related measurements. Features of interest were limited to nodes for which >50% of the data were available. Medcalc software version 16.8.4 was used to analyse the data. All tests were two-tailed and P<0.05 deemed statistically significant. Machine learning decision tree models were generated using BigML.

## Results

### Predictors of left ventricular systolic dysfunction (LVSD)

LVSD was present in 8 (10%) patients, LVEF 34 ± 6% versus 56 ± 3% in the rest of the study group, P<0.0001. Both NTproBNP and a 5-parameter A-ECG score, described previously (10, 31) had a high sensitivity, and specificity for the detection of moderate-severe LV systolic dysfunction (Table 1). The correlation between the 5-parameter A-ECG LVSD score and NTproBNP was moderate r = -0.46, 95% CI -0.64 to -0.24, P = 0.0002, suggesting some independence between the two variables.

**Table 1.** Diagnostic accuracy of Biomarkers for LVSD

| LVSD | C-statistic (95% CI) | Optimal cutpoint | Sensitivity | Specificity |
| --- | --- | --- | --- | --- |
| NTproBNP | 0.97 (0.9 to 0.99) | > 29 pg/L | 100% | 92% |
| A-ECG LVSD score | 0.97 (0.89 to 0.99) | < 0.46 | 100% | 90% |
| Singulex troponin I | 0.82 (0.71 to 0.91) | > 1.7 ng/L | 83% | 80% |
| Abbott troponin I | 0.77 (0.66 to 0.86) | > 12 ng/L | 50% | 100% |

### Predictors of left ventricular hypertrophy

28 patients (36%) had LVH defined by echocardiography based on an interventricular septum measuring ≥11mm. Median (quartile 1 and 3) interventricular septal dimension was 0.9 (0.8-1.1) cm. hs-troponin (AUC 0.85, 95% CI 0.73 to 0.92) and ECG parameters (AUC 0.75, 95% CI 0.62 to 0.85) provided accurate discrimination of patients with LVH. There was no statistically significant difference in the AUC between the three highest ranking ECG parameters identified i.e., spatial peaks QRS-T angle (by Kors), Sokolow-Lyon voltage and Cornell voltage or product. As hs-troponin was also elevated in patients with left ventricular dysfunction, it provided poor discrimination between LVH and LVSD. However NTproBNP provided good discrimination between these two disease states. Of the three ECG parameters with the highest discrimination for LVH, the spatial peaks QRS-T angle demonstrated greater informational yield than did the Sokolow Lyon criteria or Cornell Voltage/Product. Both PCA and PLS-DA demonstrated that spatial peaks QRS-T angle, spatial mean QRS-T angle, NTproBNP and hs-troponin were orthogonally separable, with Cornell voltage clustering closely with hs-troponin, and other metabolites (Figs 1 & 2). In addition, A-ECG-based discriminant analysis (28) suggested the presence of left ventricular electrical remodelling (LVER) (32-34) in 13 (17%) of the patients, most commonly in patients with either hypertension or anatomic LVH, potentially indicating intermediate pathophenotype of increased LV mass, diffuse fibrosis, or both (35). The overlap between LVER, LVH, HTN, DM and CAD within the patient population demonstrated the complex interplay between multiple disease states and A-ECG patterns (Fig. 3 and 4).

**Fig. 1.**
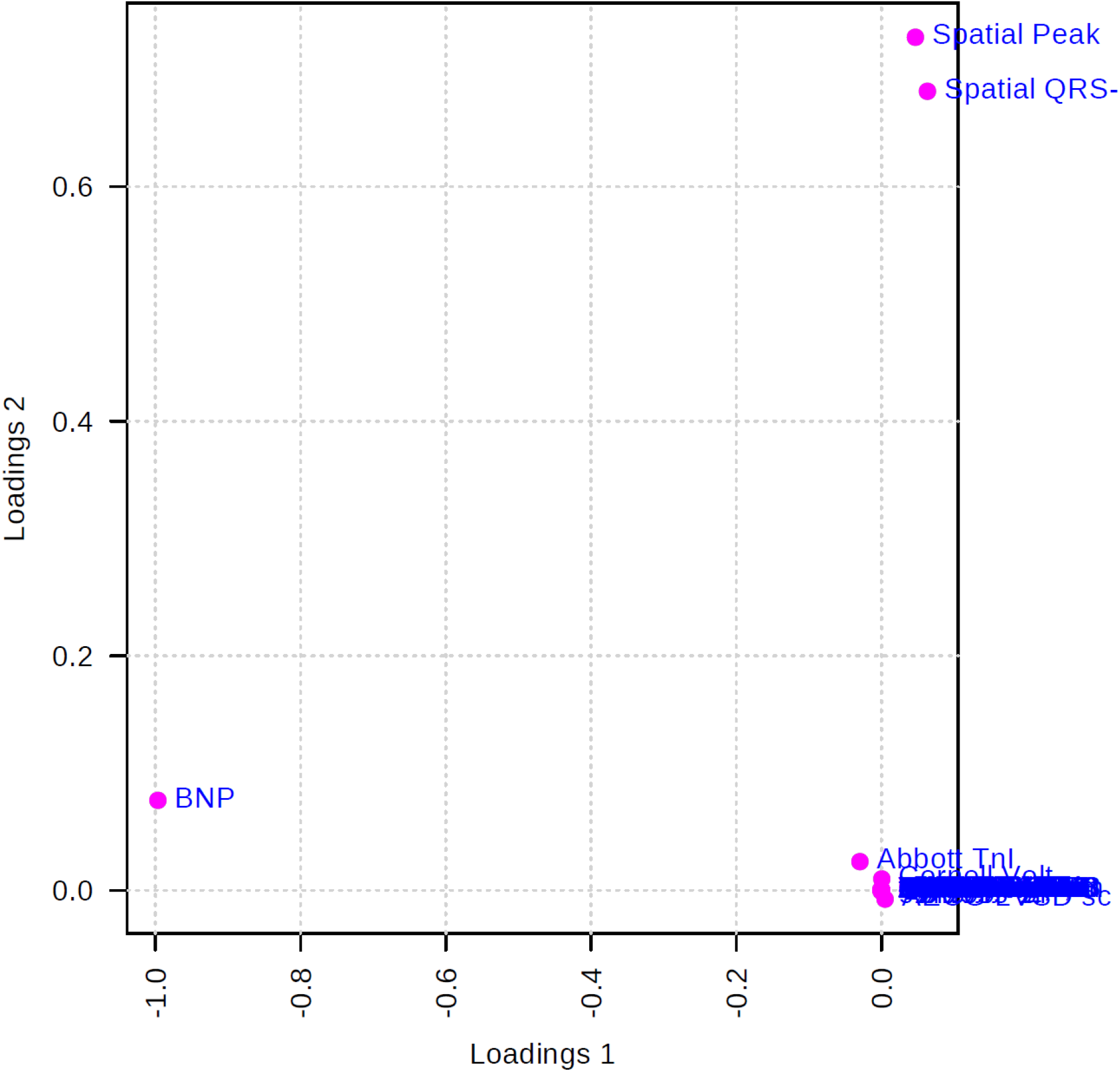
Loadings plot for the selected Principle Components in the study of left ventricular hypertrophy.

**Fig. 2.**
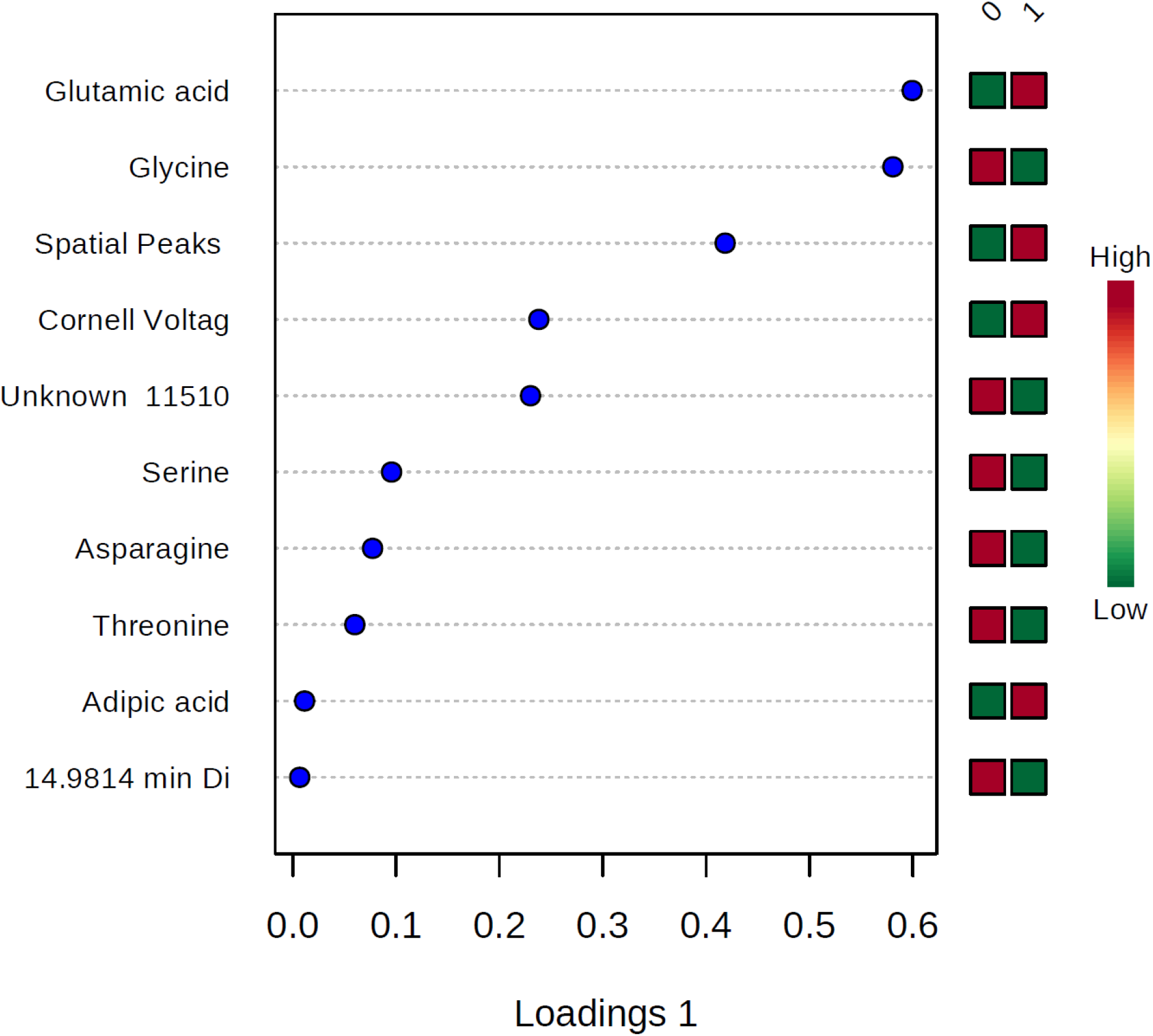
Important features identified by PLS-DA. The coloured boxes on the right indicate the relative concentrations of the corresponding metabolite in the groups without (0) and with (1) left ventricular hypertrophy, respectively.

**Figure 3.**
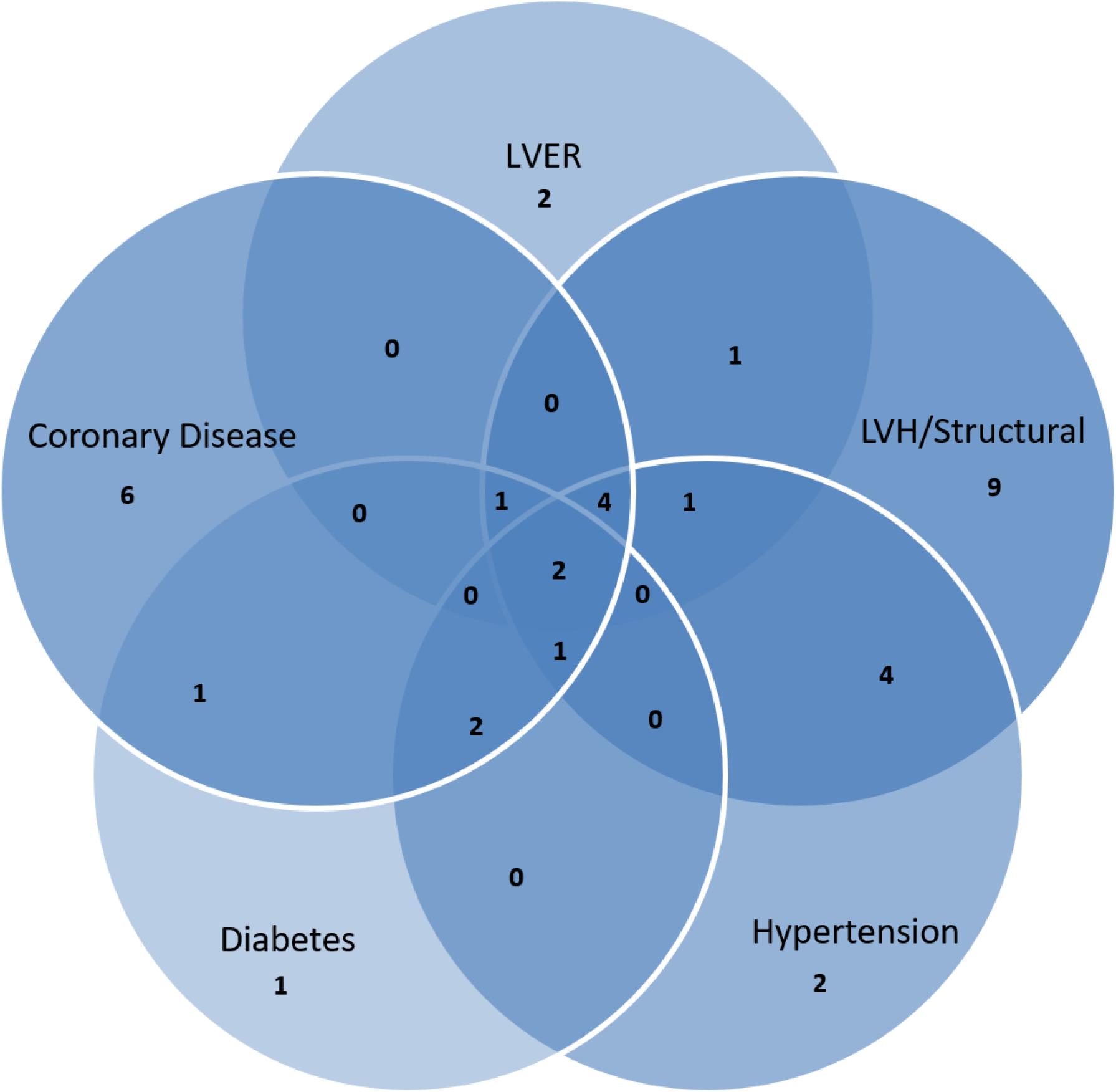
Venn diagram of varying diagnostic groups

**Figure 4.**
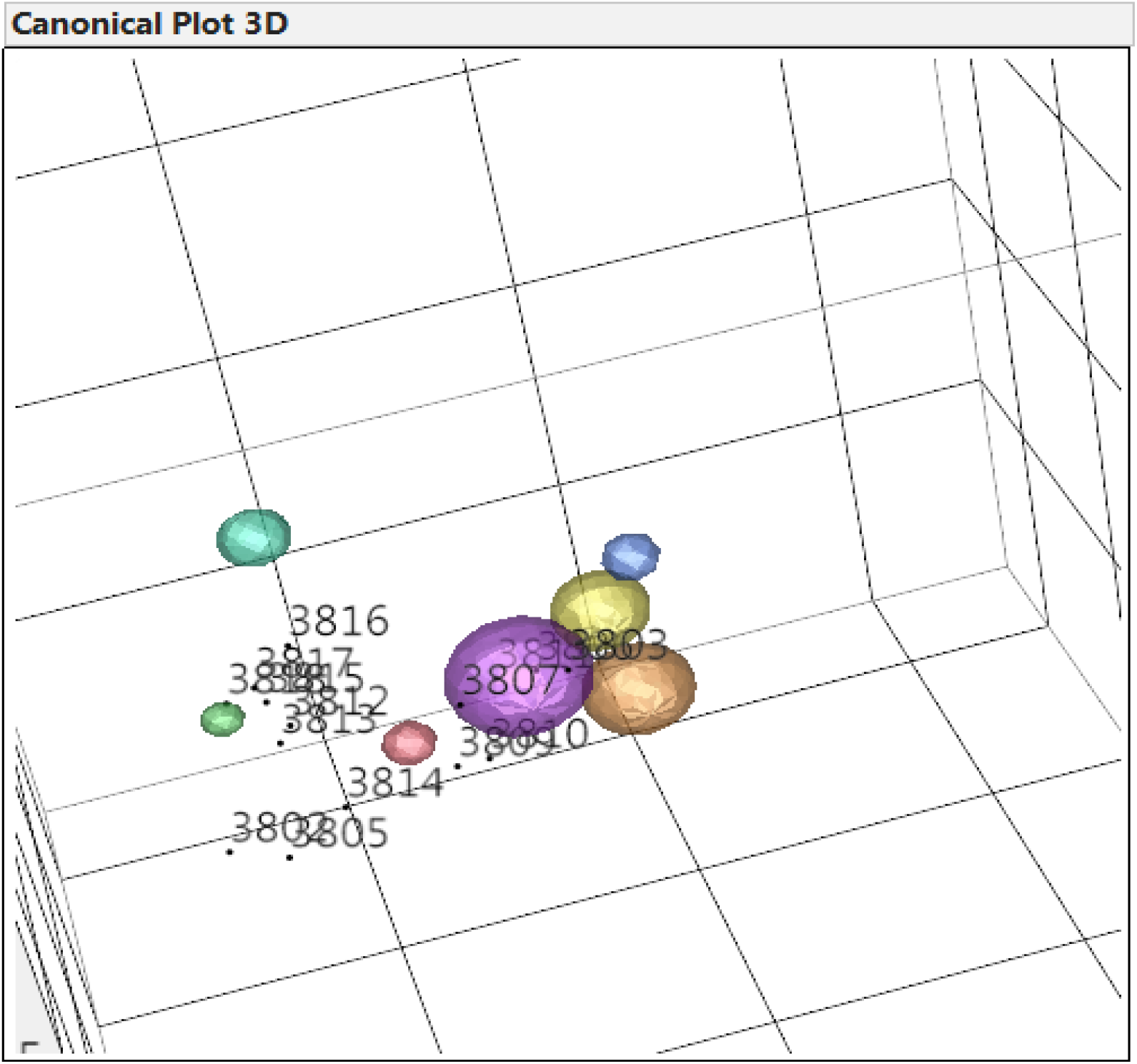
3D Canonical plot of A-ECG LDA patterns. The lime-green, red and purple spheres represent the 95% confidence intervals for the locations of groups of pre-existing patients with normal heart health, coronary artery disease and left ventricular electrical remodelling, respectively. Numbers represent certain individual patients.

### Predictors of coronary artery disease

29 (52%) of patients had CAD detected by CTCA, 9 (16%) had a stenosis ≥50% in ≥1 vessel. GLS was available on 37 (66%) of patients. GLS ≤18 had a sensitivity of 82% and specificity of 70% (AUC 0.82, 95% CI 0.66 to 0.93, p < 0.0001). eCS had a sensitivity of 100%, and specificity of 68% (AUC 0.9, 95% CI 0.79 to 0.97, p < 0.0001) in the prediction of CAD. The eCS score increased with graded severity of CAD in a dose dependent fashion (Figure 5). Mean eCS for normal versus mild or diffuse disease was 1.1 +/-1.1 versus 2.7+/- 0.7, 95% CI 1 to 2.1, p<0.0001, and for obstructive 3.7 +/-0.7, 95% CI 0.4 to 1.5, p=0.002. Both eCS ≥2 and ≥3 had an AUC 0.9, p<0.0001 for the detection of any CAD or obstructive CAD respectively. Aortic valve calcification, a subcomponent of the eCS score was also predictive of the presence of obstructive CAD with AUC 0.84, 95% CI 0.72 to 0.92, p < 0.0001. By contrast Duke and ACC/AHA clinical risk scores had AUC 0.66 and 0.53, respectively, for the same patients. In an iterative machine learning decision tree the features which together were most predictive of CAD were GLS, eCS and LA dimensions.

**Fig. 5.**
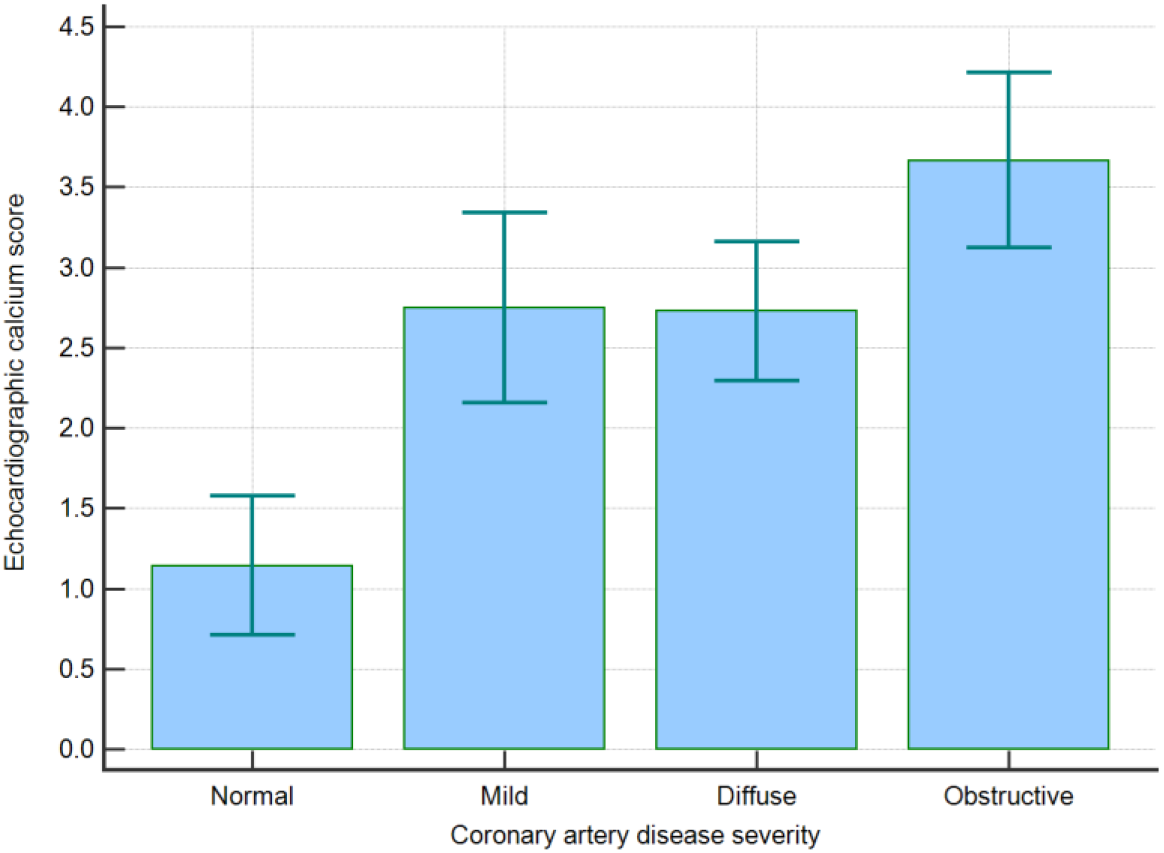
eCS score relates to severity of CAD

### Genetic risk scores

The 27 SNP genetic risk score (GRS27) did not add significant information to either the Duke or ACC/AHA clinical risk scores, nor to the eCS or GLS predictions. However there was a trend towards a higher eCS in those with a G allele in rs10455872, a SNP within LPA previously reported to be associated with aortic stenosis (2 vs 2.7, 95% CI -0.5 to 1.9, p = 0.3).(36-38) There was also a trend towards a higher GRS in those with a family history of CAD versus those without, mean GRS27 1.39 vs 1.33, 95% CI -0.03 to 0.14, p = 0.22. Mean GRS27 in this cohort without a history of acute coronary syndrome (ACS) was significantly lower than GRS27 in an ACS cohort (n=685), 1.33 vs 1.42, 95% CI 0.04 to 0.14, p = 0.0002 (Figure 6).

**Figure 6.**
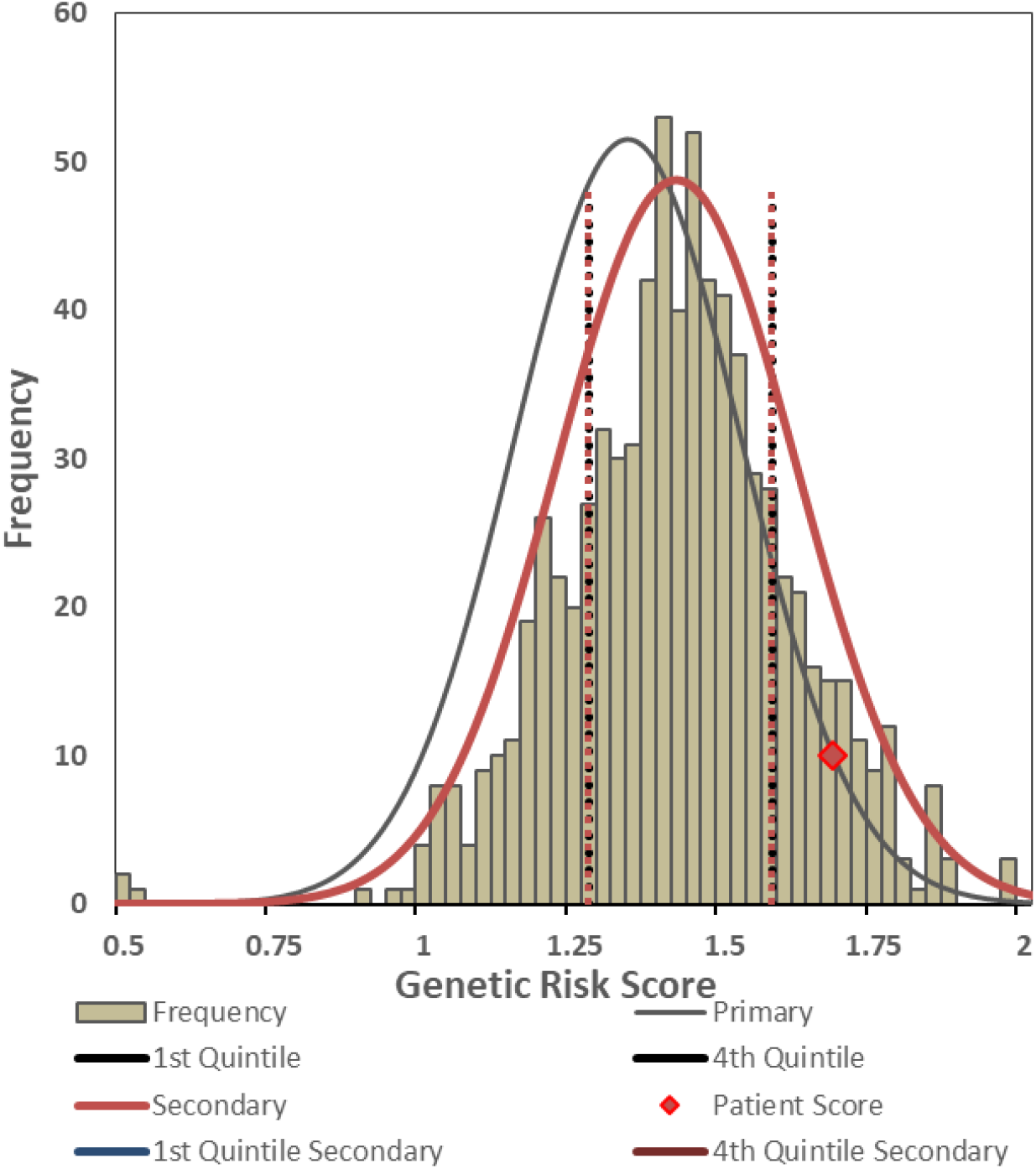
Population distribution for ACS and non-ACS patients with individual high risk GRS27 result (red diamond)

Neither rs2200733 nor rs10033464 were associated with left atrial dimensions. However an A-ECG measure of beat-by-beat variability of the P wave (27) was significantly different in rs2200733 CT or TT allele carriers. No patients with AF GRS = 0 had AF over an approximate 3 year follow-up period. 2 (7%) patients with an AF GRS ≤2 subsequently developed AF or stroke, versus 6 (16%) with a GRS ≥2, though this did not reach statistical significance. One patient, a 40 year old female with an AF GRS = 4, and a relative risk of 1.7 for AF (14), had presented with a TIA, and concurrent palpitations, though AF was not detected on inpatient telemonitoring.

### Metabolomics and Pathway Analysis

In general the difference in relative abundance of metabolite profiles varied between disease states. In order of metabolic derangements, LVSD>diabetes>hypertension>LVH>CAD with coronary artery disease having borderline to no appreciable impact on the 111 metabolites tested. Significant overlap was seen between diagnostic groups with glutamic acid being shared across diabetes, hypertension, LVH and CAD as the leading altered metabolite.

### LVSD

Important metabolites affected by the presence of LV systolic dysfunction defined by an ejection fraction <50% included malic acid, fumaric acid, cysteine, pyroglutamic acid and succinic acid. PCA and PLS-DA identified NTproBNP, A-ECG parameters spatial peaks and spatial mean QRS-T angle, and hs-troponin to be orthogonally separated. Pathway analysis indicated abnormalities in the citrate cycle (TCA cycle), alanine, aspartate and glutamate, glutathione, butanoate, phenylalanine, glyoxylate and dicarboxylate metabolism.

Using the A-ECG LVSD score < 0.5 as a definition of LVSD identified malic acid, pyroglutamic acid, cysteine, 4-hydroxyphenylactic acid and proline, glutamine, succinic, fumaric, and oxalic acid. Pathway analysis of these metabolites revealed statistically significant abnormalities in aminoacyl-tRNA biosynthesis, sulphur, taurine and hypotaurine, citric acid cycle, thiamine, pantothenate and CoA biosynthesis, pyruvate metabolism, glutathione, glycine, serine and threonine, glyoxylate and dicarboxylate, cysteine and methionine, arginine and proline metabolism, though these did not exceed the False Discovery Rate (FDR). A prior relationship between pyroglutamic acid and QTc could not be validated (39).

Using NTproBNP >35pmol/L as a biochemical definition of LVSD identified a greater number of important metabolites not only inclusive of those identified by EF <50%, and A-ECG LVSD score < 0.5, but also 4-hydroxyphenylacetic acid, cis-aconitic acid, tryptophan and malonic acid.

None of the resulting metabolites for any definition of LVSD, in univariate or multivariate testing, exceeded the diagnostic yield of NTproBNP or the A-ECG LVSD score for the diagnosis of LVSD, and it was apparent that NTproBNP, hs-troponin and A-ECG yielded similar but independent information, based on PCA and PLS-DA loadings. Combining the three definitions of LVSD in a pathway analysis (Table 2) identified statistically significant perturbations in aminoacyl-tRNA biosynthesis, nitrogen and phenylalanine metabolism, the citric acid cycle, thiamine, alanine, aspartate and glutamate metabolism. Other pathways with raw p<0.05, but in which the FDR was >0.05 also showed either biological plausibility or had been previously identified as being altered in the context of heart failure.

**Table 2.** Result from Pathway Analysis

|  | Total | Expected | Hits | Raw p | -log(p) | Holm adjust | FDR | Impact |
| --- | --- | --- | --- | --- | --- | --- | --- | --- |
| Aminoacyl-tRNA biosynthesis | 75 | 0.72 | 8 | 2.05E-07 | 1.54E+01 | 1.64E-05 | 1.64E-05 | 0.00 |
| Nitrogen metabolism | 39 | 0.37 | 5 | 2.33E-05 | 1.07E+01 | 1.84E-03 | 9.33E-04 | 0.00 |
| Phenylalanine metabolism | 45 | 0.43 | 5 | 4.77E-05 | 9.95E+00 | 3.72E-03 | 1.27E-03 | 0.06 |
| Citrate cycle (TCA cycle) | 20 | 0.19 | 3 | 7.82E-04 | 7.15E+00 | 6.02E-02 | 1.56E-02 | 0.07 |
| Thiamine metabolism | 24 | 0.23 | 3 | 1.35E-03 | 6.60E+00 | 1.03E-01 | 1.81E-02 | 0.00 |
| Alanine, aspartate and glutamate metabolism | 24 | 0.23 | 3 | 1.35E-03 | 6.60E+00 | 1.03E-01 | 1.81E-02 | 0.21 |
| Glutathione metabolism | 38 | 0.36 | 3 | 5.17E-03 | 5.26E+00 | 3.83E-01 | 5.40E-02 | 0.00 |
| Arginine and proline metabolism | 77 | 0.74 | 4 | 5.40E-03 | 5.22E+00 | 3.94E-01 | 5.40E-02 | 0.17 |
| Glycine, serine and threonine metabolism | 48 | 0.46 | 3 | 9.96E-03 | 4.61E+00 | 7.17E-01 | 8.86E-02 | 0.19 |
| Glyoxylate and dicarboxylate metabolism | 50 | 0.48 | 3 | 1.12E-02 | 4.50E+00 | 7.92E-01 | 8.92E-02 | 0.15 |
| Phenylalanine, tyrosine and tryptophan biosynthesis | 27 | 0.26 | 2 | 2.65E-02 | 3.63E+00 | 1.00E+00 | 1.89E-01 | 0.01 |
| beta-Alanine metabolism | 28 | 0.27 | 2 | 2.84E-02 | 3.56E+00 | 1.00E+00 | 1.89E-01 | 0.01 |
| Tyrosine metabolism | 76 | 0.73 | 3 | 3.40E-02 | 3.38E+00 | 1.00E+00 | 2.08E-01 | 0.05 |
| Pyruvate metabolism | 32 | 0.31 | 2 | 3.64E-02 | 3.31E+00 | 1.00E+00 | 2.08E-01 | 0.14 |
| Propanoate metabolism | 35 | 0.33 | 2 | 4.29E-02 | 3.15E+00 | 1.00E+00 | 2.29E-01 | 0.00 |
| Butanoate metabolism | 40 | 0.38 | 2 | 5.47E-02 | 2.91E+00 | 1.00E+00 | 2.73E-01 | 0.04 |
| Fatty acid biosynthesis | 49 | 0.47 | 2 | 7.83E-02 | 2.55E+00 | 1.00E+00 | 3.69E-01 | 0.00 |
| D-Glutamine and D-glutamate metabolism | 11 | 0.11 | 1 | 1.00E-01 | 2.30E+00 | 1.00E+00 | 4.46E-01 | 0.03 |
| Pyrimidine metabolism | 60 | 0.57 | 2 | 1.11E-01 | 2.20E+00 | 1.00E+00 | 4.66E-01 | 0.00 |
| Linoleic acid metabolism | 15 | 0.14 | 1 | 1.35E-01 | 2.01E+00 | 1.00E+00 | 5.38E-01 | 0.00 |
| Cyanoamino acid metabolism | 16 | 0.15 | 1 | 1.43E-01 | 1.95E+00 | 1.00E+00 | 5.44E-01 | 0.00 |
| Sulfur metabolism | 18 | 0.17 | 1 | 1.59E-01 | 1.84E+00 | 1.00E+00 | 5.79E-01 | 0.03 |
| Taurine and hypotaurine metabolism | 20 | 0.19 | 1 | 1.75E-01 | 1.74E+00 | 1.00E+00 | 6.10E-01 | 0.00 |
| Purine metabolism | 92 | 0.88 | 2 | 2.19E-01 | 1.52E+00 | 1.00E+00 | 7.06E-01 | 0.00 |
| Pantothenate and CoA biosynthesis | 27 | 0.26 | 1 | 2.29E-01 | 1.47E+00 | 1.00E+00 | 7.06E-01 | 0.00 |
| Valine, leucine and isoleucine biosynthesis | 27 | 0.26 | 1 | 2.29E-01 | 1.47E+00 | 1.00E+00 | 7.06E-01 | 0.01 |
| Glycolysis or Gluconeogenesis | 31 | 0.30 | 1 | 2.59E-01 | 1.35E+00 | 1.00E+00 | 7.67E-01 | 0.00 |
| Methane metabolism | 34 | 0.32 | 1 | 2.80E-01 | 1.27E+00 | 1.00E+00 | 8.00E-01 | 0.00 |
| Ubiquinone and other terpenoid-quinone biosynthesis | 36 | 0.34 | 1 | 2.94E-01 | 1.22E+00 | 1.00E+00 | 8.11E-01 | 0.00 |
| Valine, leucine and isoleucine degradation | 40 | 0.38 | 1 | 3.21E-01 | 1.14E+00 | 1.00E+00 | 8.56E-01 | 0.02 |
| Nicotinate and nicotinamide metabolism | 44 | 0.42 | 1 | 3.47E-01 | 1.06E+00 | 1.00E+00 | 8.61E-01 | 0.00 |
| Histidine metabolism | 44 | 0.42 | 1 | 3.47E-01 | 1.06E+00 | 1.00E+00 | 8.61E-01 | 0.14 |
| Lysine degradation | 47 | 0.45 | 1 | 3.66E-01 | 1.01E+00 | 1.00E+00 | 8.61E-01 | 0.00 |
| Primary bile acid biosynthesis | 47 | 0.45 | 1 | 3.66E-01 | 1.01E+00 | 1.00E+00 | 8.61E-01 | 0.01 |
| Cysteine and methionine metabolism | 56 | 0.54 | 1 | 4.20E-01 | 8.69E-01 | 1.00E+00 | 9.59E-01 | 0.13 |
| Tryptophan metabolism | 79 | 0.75 | 1 | 5.38E-01 | 6.21E-01 | 1.00E+00 | 1.00E+00 | 0.11 |
| Porphyrin and chlorophyll metabolism | 104 | 0.99 | 1 | 6.40E-01 | 4.47E-01 | 1.00E+00 | 1.00E+00 | 0.00 |

Raw p is the original p value calculated from the enrichment analysis; the Holm p is the p value adjusted by Holm-Bonferroni method; the FDR p is the p value adjusted using False Discovery Rate; the Impact is the pathway impact value calculated from pathway topology analysis.

### Diabetes

7 (9%) patients had type II diabetes. Benzoic acid, glutamic acid, 2-hydroxybutyric acid, fumaric acid and valine were increased in those with diabetes, though after multiplicity correction only benzoic acid remained statistically significant. Pathways perturbed included several similar to heart failure but notably included butanoate and propanoate metabolism though these did not reach FDR.

### Hypertension and LVH

Glutamic acid, lactic acid, ethyl 6-methoxy-3-quinolinylcarbamate (NIST), ornithine, succinic acid, malic acid, proline, pyruvic acid, pyroglutamic acid, glycine were altered in those with hypertension with expected overlap with metabolites altered in those with LVH, i.e., glutamic acid, glycine, serine, asparagine, threonine, adipic acid. However after correction for multiplicity only glutamic acid, lactic acid, ethyl 6-methoxy-3-quinolinylcarbamate NIST, ornithine, succinic acid and malic acid were altered. These indicated abnormalities in the citrate cycle (TCA cycle),alanine, aspartate and glutamate, pyruvate, propanoate, glutathione, butanoate, glyoxylate and dicarboxylate metabolism pathways.

### Coronary artery disease

Glutamic acid, trans-4-hydroxyproline, DPA (C22_5n-3,6,9,12,15c), alanine, Ethyl 6-methoxy-3-quinolinylcarbamate (NIST), pyruvic acid and lactic acid appeared altered in those with coronary artery disease. However none of these reached FDR. In patients in whom the GRS pathway indicated risk other than lipids, the metabolites tryptophan and ornithine featured but did not reach FDR.

#### Global interactome

Network features in the global interactome included notable correlations between measures of heart rate variability and caffeine, eicosadecanoic and eicosapentaenoic acid (Figure 7). LA volume index had a high index of betweenness centrality, connecting metabolomics, A-ECG, echocardiography and protein biomarkers supporting the axiom that the left atrium serves as a barometer of the heart (Figure 8).(40) The network interactome can be visualised here https://theranosticslab.bitbucket.io/network_tsne/?datafiles=myHealth-Tests.json,myHealth-Patients.json

**Figure 7.**
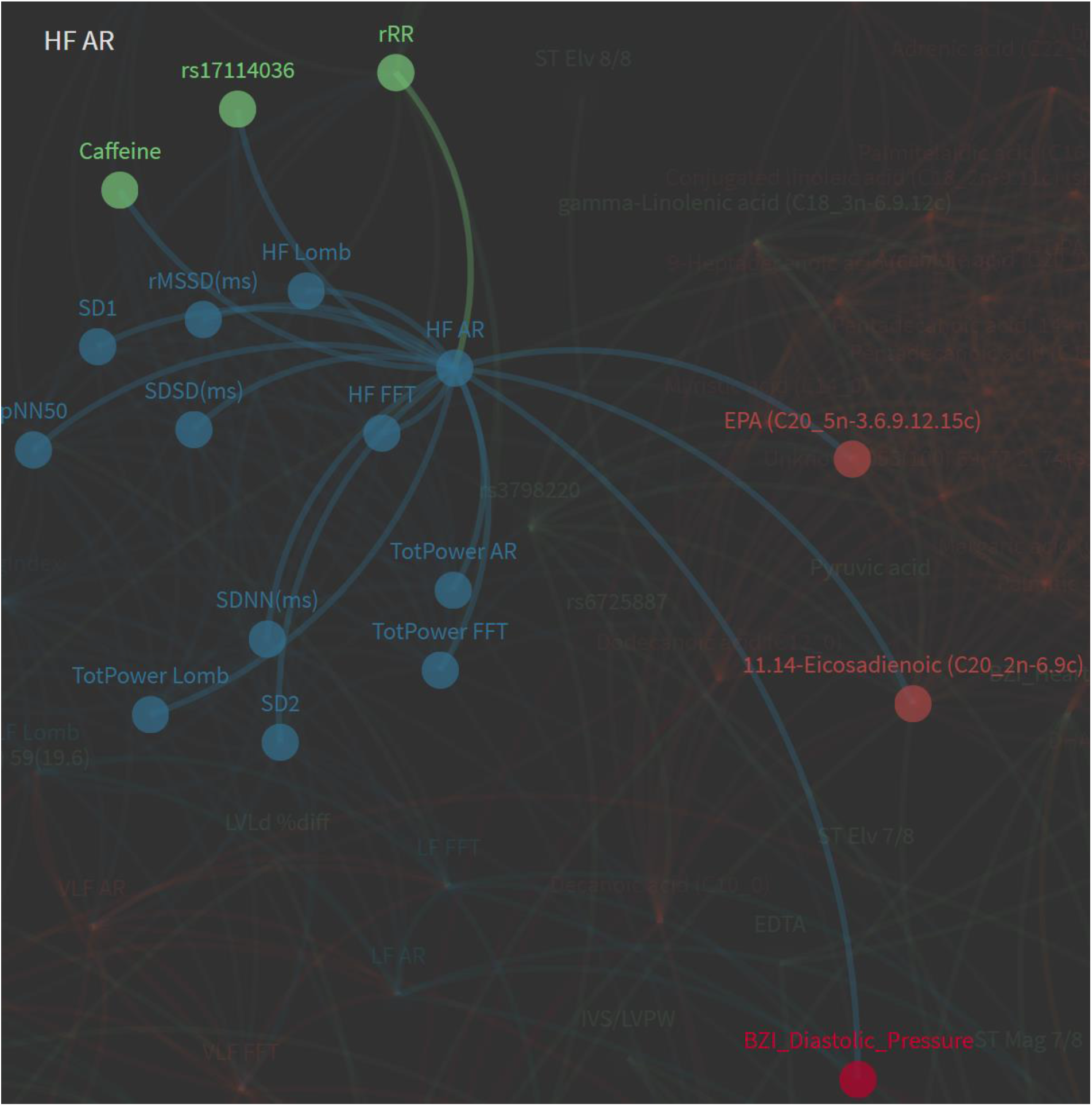
Metabolomic and nutritional relationships with Heart Rate Variability (HRV)

**Figure 8.**
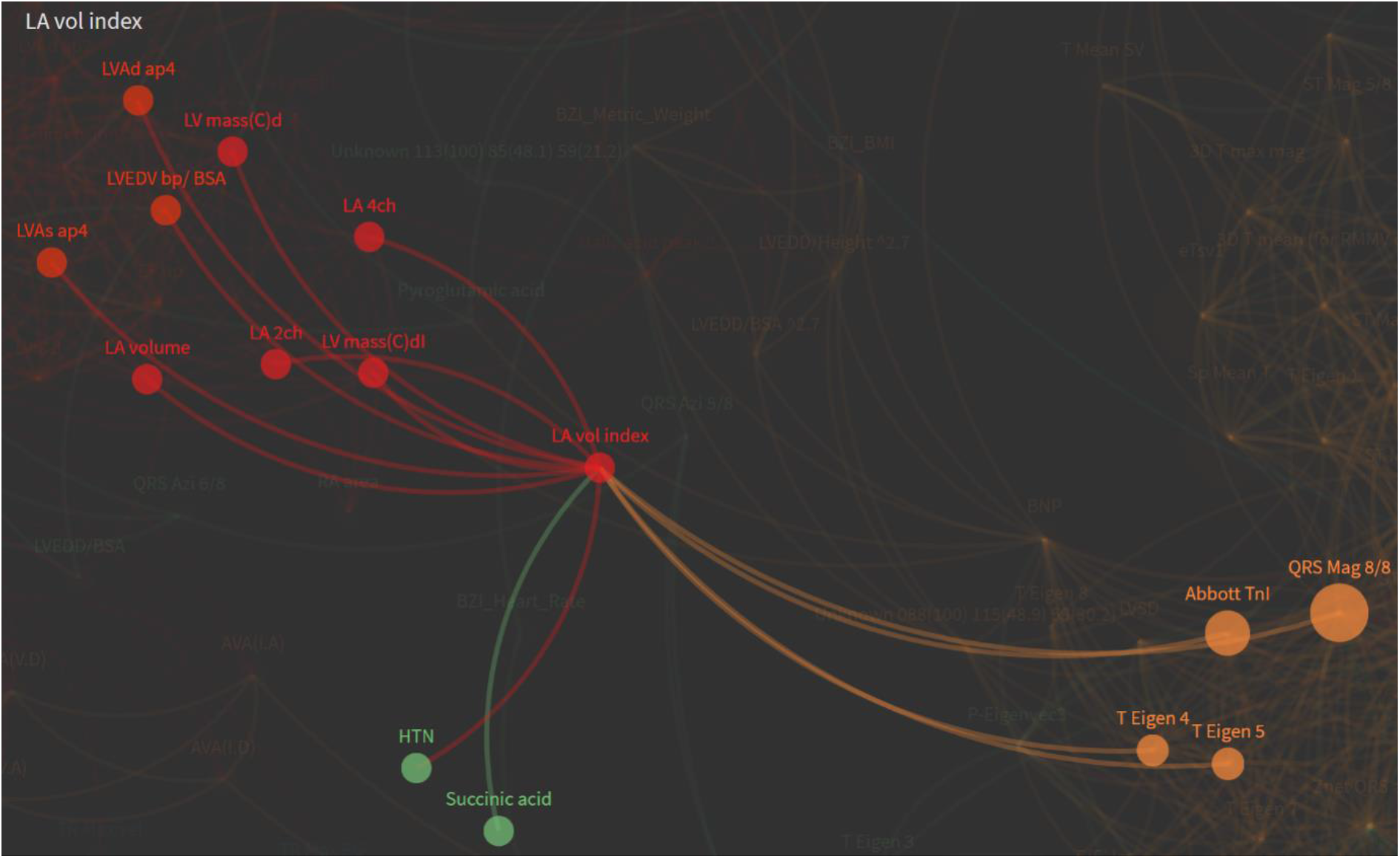
LA volume index demonstrates high degree of betweenness centrality across multiple domains

## Discussion

In this pilot study we demonstrated a number of interesting findings that warrant further investigation. Firstly we showed the potential research and clinical utility of combining multidimensional data spanning genetic, metabolomic, protein biomarkers, advanced ECG (spatial and temporal) and cardiac imaging data in the same patients. This data showed both diagnostic and prognostic capability. Although this study tested multiple hypotheses, even after correction for multiplicity several results remained statistically significant. Furthermore several of these findings have already been demonstrated to have potential clinical utility in other studies. We essentially aimed to gather data from existing clinical tools, such as ECG and echocardiography to discover the bare minimum dataset that would yield the maximal diagnostic yield for a broad array of cardiac diagnoses.

Validating an earlier finding we have shown that a 5-parameter A-ECG score has high discriminatory accuracy for LV systolic dysfunction, in those with moderate LV systolic dysfunction. This was comparable, though somewhat independent to NTproBNP. As digital ECG is a commonly performed in patients with suspected cardiac disease, the addition of an A-ECG, essentially a software algorithm, could further improve diagnoses in an ambulatory clinic. Conventional ECG screening of general populations is currently considered not cost-effective and is therefore not recommended by cardiology guidelines however the enhancement of ECG with advanced parameters and machine learning may require a rethink of that position, as more population data supporting A-ECG becomes available (41). Although the c-statistic for the diagnosis of LVSD was high for both NTproBNP and A-ECG it is important to note that the mean LVEF in this population was 34% and discrimination may not be as accurate in patients with mild systolic dysfunction. However it is possible that these two biomarkers may play an adjunctive role, as A-ECG includes additional multidimensional prognostic data, such as markers of arrhythmic risk (10). It is unsurprising that A-ECG and metabolomics appear closely inter-related and in this study we identified a metabolite profile for a novel A-ECG score for LVSD, which we have previously shown to correlate with GLS, heart failure outcomes and ventricular arrhythmia (10).

By integrating multiple sources of data including metabolomics we have also demonstrated the potential for these methods to redefine disease states. Heart failure for instance is a clinical diagnosis, which is commonly subtyped into heart failure with preserved ejection fraction (HFpEF) and reduced ejection fraction (HFrEF). Although NTproBNP is an effective diagnostic and prognostic biomarker in HF it is less capable of identifying HFpEF than HFrEF (42). Whilst LVEF is the sine qua non used to distinguish these two conditions, metabolomics has also been used to demonstrate their differing underlying molecular mechanisms (43). Metabolomics has also been shown to have a better prognostic role than NTproBNP and could be used as a discriminator for CRT or ICD device therapy if further validation occurs (44-46). Similarly we have previously shown that A-ECG, akin to the ‘cardioelectrome’, has a similar capacity to subtype HF patients and identify ICD candidates (10). In this study we did could not show significant diagnostic yield for GC-MS metabolomics, however that was most likely due to the limited range of metabolites tested, rather than a failure of the technology per se. Whilst it is clear that use of LVEF alone has deficiencies in defining the syndrome of heart failure, it is not yet clear whether an ‘omic method could supersede or augment existing diagnostic tools. What this study showed is that metabolomics is clearly a powerful tool capable of demonstrating distinct differences between small numbers of patients with disease and of identifying potential mechanistic pathways, e.g., mitochondrial energetic metabolism, which could become treatment targets. The most notable of these pathways in this study was thiamine metabolism. Prior observational literature has shown that chronic furosemide administration results in thiamine deficiency (47), and small randomized trials have proposed that thiamine supplementation has a role in HF treatment (48, 49). Pharmacological and nutritional studies including metabolomics, as a method of monitoring treatment response, are few but this could result in smaller trials, new surrogate outcomes to supplant clinical ones and could contribute insights into the efficacy and systems pharmacology of alternative therapies (50-52).

Although large scale population screening studies of metabolomics have not yet been shown to exceed standard clinical HF risk tools, they are giving insights into potential mechanisms of disease, e.g., a recent finding identifying phenylalanine metabolism (53). Metabolomics is highly amenable to population screening, due to its exceedingly low cost per test. High throughput metabolomics is already used in newborn screening programs for inborn errors of metabolism (IEMs).

Almost certainly the current diagnostic strategies and schema of heart failure will need to change once larger integrated ‘omic datasets are available, including genomic sequence data and broad clinical metadata (54).This is likely to include the newly discovered Titin truncating variants, present in between 10-20% of HF patients and relevant with respect to both cardiac metabolism (55) and arrhythmic outcomes (56).

Further to the redefinition of disease processes in this study we identified a relatively new endophenotype of left ventricular hypertrophy, i.e. left ventricular electrical remodeling (LVER) (32-34). Whilst fairly interdependent this A-ECG finding was also sometimes present in patients without discernible LVH, some with a history of recurrent chest pain admissions. Further work will be required to address causality, though since LVH is associated with cardiac chest pain it would seem probable that LVER could be an important contributor to chest pain syndromes. With ECG being clinically more available than echocardiography in patients with chest pain, this could provide a useful clinical adjunct to standard history and hs-troponin testing. We were unable to demonstrate the ability of hs-troponin to exclude obstructive coronary disease in this cohort as shown by Lee et al (57), due to the high prevalence of LVH. However we did validate that hs-troponin has an important role in identifying patients in increased LV mass (58). A-ECG also provided some independent diagnostic information regarding LV mass, which could potentially be augmented by both hs-troponin and metabolomic data (e.g. glycine and glutamate), however greater power is required to be certain of this finding. It is noteworthy that an inborn error of glycine metabolism has been described which results in LVH, supporting a causal role.(59)

Significant effort has been placed in the identification of metabolomic markers of cardiovascular disease presence or risk (60-63). Whilst some improvement in net reclassification of CVD risk has been achieved with metabolomics the increased yield has not yet translated metabolomics into clinical care. This may in part be because metabolomics has a greater role in identifying the disease processes causative of cardiovascular disease, such as diabetes and hypertension, rather than coronary disease itself. The latter may not be particularly metabolic, especially when defined anatomically. Caution has also been raised about replication of metabolomic signatures in real world settings, as signatures for multiple conditions frequently overlap (64), something we also demonstrated in this study using other biomarkers. Of all the resting investigations we used, the greatest yield for diagnosis of anatomically-defined CAD came from echocardiography, a technology which is now highly portable, and becoming more available and inexpensive.(65) Although resting echocardiography has not traditionally been used for screening patients for CAD, we have validated a potential role of advanced echocardiography i.e. eCS score and GLS to identify those with CAD (6, 66-68). Integrating other forms of data such as hs-troponin, NTproBNP, and genetic risk scores may increase the yield and predictive ability of advanced echocardiography, however in this small cohort we could not demonstrate this definitively, due to the poor individual predictive capacity of GRS27. It is worth noting that the CAD GRS27(13) has rapidly been superseded by GRS50 (69), GRS49×10^3^ (70) and more recently GRS 1.7×10^6^ (71), which has been shown to carry a risk equivalent to monogenic causes of CAD, such as FH mutations (72). As a GRS has a role in risk prediction in younger patients it may potentially augment eCS. As the current CAD GRS contains information (i.e. LPA SNPs) relevant to aortic stenosis (37) it also seems likely that genomic risk prediction and echocardiography will see further convergence. Technologically this convergence is already being seen in the evolution of capacitive micromachined ultrasonic (CMUT) transducers, used in flat panel wearable ultrasound and complementary metal– oxide–semiconductor (CMOS) sequencing technology (73, 74).

Since eCS has many similarities to coronary artery calcium scoring (CAC), often performed by automated analysis of computed tomography images, it would seem likely that the eCS score would lend itself well to machine vision applications in echocardiography (75). This could be further supplemented with other echocardiographic data such as GLS, chamber dimensions, and eventually multi-omic data. For this reason we have been piloting a novel 5-min echocardiography protocol in our hospital, aimed at capturing the minimal data set required for machine vision interpretation (76).

A GRS for AF demonstrated relatively poor ability to predict incident atrial fibrillation in follow up, though this is to be expected with a panel containing few SNPs. Similar to the GRS for CAD the number of GWAS AF SNPs, in ever larger cohorts, has been growing rapidly (77). We were unable to demonstrate previous findings of an association between rs2200733 and LA dimensions (78), however we did show a novel association between this SNP and an A-ECG parameter of atrial activity. rs2200733 has been shown to have predictive capacity in the context of not only DC cardioversion and pulmonary vein isolation (79, 80), but also ischaemic stroke (22). It is therefore interesting that one patient with a moderate AF GRS risk also presented with a TIA.

Using a network analysis, temporal A-ECG data demonstrated interesting relationships between measures of HRV, plasma DHA (n-3) and EPA (n-6) fatty acids as well as caffeine. The relationship between the nutritional biomarkers (DHA/EPA) and heart rate variability have previously been described in patients with increased cardiovascular risk, and is modifiable by nutritional supplementation (81). Similarly caffeine is known to alter HRV in both healthy and diabetic patients (82, 83), but not in patients with heart failure (84). With the widespread availability of wearable devices measuring HRV it would therefore seem logical that nutritional and health status could be probed by consumer wearables in the community (2). Lim et al recently evaluated the output of wearable devices as they related to cardiac imaging with cardiac magnetic resonance imaging and lipidomics, demonstrating a utility in wearables predicting LV remodeling (5). To expand upon these concepts further we are currently evaluating the utility of a Virtual Reality (VR) mental stress test, A-ECG metabolomics and wearables in patients with heart failure (85).

## Conclusion

We have demonstrated in this study a relatively accurate multi-omic approach to screening for common cardiovascular diseases that can be achieved at relatively low cost. Whilst this pilot project was rudimentary compared to larger, ongoing personal integrated ‘omic programs, real time integration of such programs will require considerable research effort and cost to achieve. To handle the high computational demand, such programs will also require Cloud computing, connected to the graphic and tensor processing unit clusters theoretically ideal for advanced image processing. And for clinical purposes such infrastructure also carries with it notable privacy and security issues. We have herein instead shown that a semi-targeted approach, using best available knowledge, may not only be useful, but also simpler to clinically implement at a local level. However we have begun work on a virtual machine generating both data-driven and computational biophysical models to back propagate missing clinical data, due its sparseness, and address the problems associated with causal inference (86).

Because many of the described technologies already use readily available clinical data, rather than simply stating that more research is required, it would seem plausible that they could be employed in clinical care and evaluated in practice in a constant audit with iterative feedback and adaptation, i.e., rather than delaying adoption potentially for decades while awaiting full clinical validation. Several interventional technologies have been adopted into clinical care with lesser evidence and still persist, even after randomized sham trials have shown limited or no benefit. Although some barriers such as methods standardization exist, further delay in the clinical implementation of these useful technologies may not be justifiable. We would suggest that a process of rapid adoption with constant monitoring and iterative adaptation could achieve a significant gain over the current status quo. Intervention studies, randomized trials and mass consortia data warehouses are still required but could be embedded into electronic health records or registries to be more effective (87).

## Limitations

There are several limitations to this study, mainly relating to the small sample size and testing of multiple hypothesis with an increased chance of a type I error. However where possible we have used an adjustment for multiplicity with a measure of the false discovery rate (FDR). Furthermore we have limited the analyses to diagnostic methods which already have a literature supporting their use.

## Data Availability

Data is available on request

## References

1. Guo L, Milburn MV, Ryals JA, Lonergan SC, Mitchell MW, Wulff JE, et al. Plasma metabolomic profiles enhance precision medicine for volunteers of normal health. Proc Natl Acad Sci U S A. 2015;112(35):E4901–10.

2. Li X, Dunn J, Salins D, Zhou G, Zhou W, Schussler-Fiorenza Rose SM, et al. Digital Health: Tracking Physiomes and Activity Using Wearable Biosensors Reveals Useful Health-Related Information. PLoS biology. 2017;15(1):e2001402.

3. Price ND, Magis AT, Earls JC, Glusman G, Levy R, Lausted C, et al. A wellness study of 108 individuals using personal, dense, dynamic data clouds. Nature biotechnology. 2017.

4. Piening BD, Zhou W, Contrepois K, Rost H, Gu Urban GJ, Mishra T, et al. Integrative Personal Omics Profiles during Periods of Weight Gain and Loss. Cell systems. 2018.

5. Lim WK, Davila S, Teo JX, Yang C, Pua CJ, Blöcker C, et al. Beyond fitness tracking: The use of consumer-grade wearable data from normal volunteers in cardiovascular and lipidomics research. PLoS Biology. 2018;16(2):e2004285.

6. Corciu AI, Siciliano V, Poggianti E, Petersen C, Venneri L, Picano E. Cardiac calcification by transthoracic echocardiography in patients with known or suspected coronary artery disease. International journal of cardiology. 2010;142(3):288–95.

7. Gaibazzi N, Baldari C, Faggiano P, Albertini L, Faden G, Pigazzani F, et al. Cardiac calcium score on 2D echo: correlations with cardiac and coronary calcium at multi-detector computed tomography. Cardiovascular ultrasound. 2014;12:43.

8. Gaibazzi N, Porter TR, Agricola E, Cioffi G, Mazzone C, Lorenzoni V, et al. Prognostic value of echocardiographic calcium score in patients with a clinical indication for stress echocardiography. JACC Cardiovascular imaging. 2015;8(4):389–96.

9. Schlegel TT, Kulecz WB, Feiveson AH, Greco EC, DePalma JL, Starc V, et al. Accuracy of advanced versus strictly conventional 12-lead ECG for detection and screening of coronary artery disease, left ventricular hypertrophy and left ventricular systolic dysfunction. BMC Cardiovasc Disord. 2010;10:28.

10. Gleeson S, Liao YW, Dugo C, Cave A, Zhou L, Ayar Z, et al. ECG-derived spatial QRS-T angle is associated with ICD implantation, mortality and heart failure admissions in patients with LV systolic dysfunction. PloS one. 2017;12(3):e0171069.

11. Cortez D, Schlegel TT, Ackerman MJ, Bos JM. ECG-derived spatial QRS-T angle is strongly associated with hypertrophic cardiomyopathy. J Electrocardiol. 2017;50(2):195–202.

12. Wang T, Dugo C, Whalley G, Wynne Y, Semple H, Smith K, et al. Diagnostic Utility of High Sensitivity Troponins for Echocardiographic Markers of Structural Heart Disease. Medical Sciences. 2018;6(1):17.

13. Mega JL, Stitziel NO, Smith JG, Chasman DI, Caulfield M, Devlin JJ, et al. Genetic risk, coronary heart disease events, and the clinical benefit of statin therapy: an analysis of primary and secondary prevention trials. Lancet. 2015;385(9984):2264–71.

14. Lubitz SA, Sinner MF, Lunetta KL, Makino S, Pfeufer A, Rahman R, et al. Independent susceptibility markers for atrial fibrillation on chromosome 4q25. Circulation. 2010;122(10):976–84.

15. Muse ED, Wineinger NE, Spencer EG, Peters M, Henderson R, Zhang Y, et al. Validation of a genetic risk score for atrial fibrillation: A prospective multicenter cohort study. PLOS Medicine. 2018;15(3):e1002525.

16. Sun L, Tian L, Xu J, Zhang Z, Liu X. Chromosome 4q25 Variants and Age at Onset of Ischemic Stroke. Molecular neurobiology. 2017;54(5):3388–94.

17. Sun L, Zhang Z, Xu J, Xu G, Liu X. Chromosome 4q25 Variants rs2200733, rs10033464, and rs1906591 Contribute to Ischemic Stroke Risk. Molecular neurobiology. 2016;53(6):3882–90.

18. Su L, Shen T, Xie J, Yan Y, Chen Z, Wu Y, et al. Association of GWAS-Supported Variants rs2200733 and rs6843082 on Chromosome 4q25 with Ischemic Stroke in the Southern Chinese Han Population. Journal of molecular neuroscience : MN. 2015;56(3):585–92.

19. Cao YY, Ma F, Wang Y, Wang DW, Ding H. Rs2200733 and rs10033464 on chromosome 4q25 confer risk of cardioembolic stroke: an updated meta-analysis. Mol Biol Rep. 2013;40(10):5977–85.

20. Wnuk M, Pera J, Jagiella J, Szczygiel E, Ferens A, Spisak K, et al. The rs2200733 variant on chromosome 4q25 is a risk factor for cardioembolic stroke related to atrial fibrillation in Polish patients. Neurologia i neurochirurgia polska. 2011;45(2):148–52.

21. Shi L, Li C, Wang C, Xia Y, Wu G, Wang F, et al. Assessment of association of rs2200733 on chromosome 4q25 with atrial fibrillation and ischemic stroke in a Chinese Han population. Human genetics. 2009;126(6):843–9.

22. Gretarsdottir S, Thorleifsson G, Manolescu A, Styrkarsdottir U, Helgadottir A, Gschwendtner A, et al. Risk variants for atrial fibrillation on chromosome 4q25 associate with ischemic stroke. Annals of neurology. 2008;64(4):402–9.

23. Holley A, Matsis K, Northcott H, Gladding P, Harding S, Larsen P. Genetic Risk Scoring in a Young Myocardial Infarction (MI) Population. Heart, Lung and Circulation.25:S283–S4.

24. Batdorf BH, Feiveson AH, Schlegel TT. The effect of signal averaging on the reproducibility and reliability of measures of T-wave morphology. Journal of electrocardiology. 2006;39(3):266–70.

25. Kors JA, van Herpen G, Sittig AC, van Bemmel JH. Reconstruction of the Frank vectorcardiogram from standard electrocardiographic leads: diagnostic comparison of different methods. European heart journal. 1990;11(12):1083–92.

26. Rautaharju PM, Kooperberg C, Larson JC, LaCroix A. Electrocardiographic predictors of incident congestive heart failure and all-cause mortality in postmenopausal women: the Women’s Health Initiative. Circulation. 2006;113(4):481–9.

27. Starc V, Schlegel TT. Real-time multichannel system for beat-to-beat QT interval variability. J Electrocardiol. 2006;39(4):358–67.

28. Gladding P, Cave A, Zareian M, Smith K, Hussan J, Hunter P, et al. Open Access Integrated Therapeutic and Diagnostic Platforms for Personalized Cardiovascular Medicine. Journal of Personalized Medicine. 2013;3(3):203.

29. Gladding PA, Cave A, Zareian M, Smith K, Hussan J, Hunter P, et al. Open access integrated therapeutic and diagnostic platforms for personalized cardiovascular medicine. J Pers Med. 2013;3(3):203–37.

30. Smart KF, Aggio RB, Van Houtte JR, Villas-Boas SG. Analytical platform for metabolome analysis of microbial cells using methyl chloroformate derivatization followed by gas chromatography-mass spectrometry. Nat Protoc. 2010;5(10):1709–29.

31. Johnson K, Neilson S, To A, Amir N, Cave A, Scott T, et al. Advanced Electrocardiography Identifies Left Ventricular Systolic Dysfunction in Non-Ischemic Cardiomyopathy and Tracks Serial Change over Time. J Cardiovasc Dev Dis. 2015;2(2):93–107.

32. Cutler MJ, Jeyaraj D, Rosenbaum DS. Cardiac electrical remodeling in health and disease. Trends in pharmacological sciences. 2011;32(3):174–80.

33. Rodriguez-Padial L, Bacharova L. Electrical remodeling in left ventricular hypertrophy—is there a unifying hypothesis for the variety of electrocardiographic criteria for the diagnosis of left ventricular hypertrophy? Journal of Electrocardiology. 2012;45(5):494–7.

34. Estes EH, Jr. ECG manifestations of left ventricular electrical remodeling. J Electrocardiol. 2012;45(6):612–6.

35. Maanja M, Wieslander B, Schlegel TT, Bacharova L, Abu Daya H, Fridman Y, et al. Diffuse Myocardial Fibrosis Reduces Electrocardiographic Voltage Measures of Left Ventricular Hypertrophy Independent of Left Ventricular Mass. J Am Heart Assoc. 2017;6(1).

36. Kamstrup PR, Tybjærg-Hansen A, Nordestgaard BG. Elevated Lipoprotein(a) and Risk of Aortic Valve Stenosis in the General Population. Journal of the American College of Cardiology. 2014;63(5):470–7.

37. Cairns BJ, Coffey S, Travis RC, Prendergast B, Green J, Engert JC, et al. A Replicated, Genome-Wide Significant Association of Aortic Stenosis With a Genetic Variant for Lipoprotein(a). Meta-Analysis of Published and Novel Data. 2017;135(12):1181–3.

38. Thanassoulis G, Campbell CY, Owens DS, Smith JG, Smith AV, Peloso GM, et al. Genetic Associations with Valvular Calcification and Aortic Stenosis. The New England journal of medicine. 2013;368(6):503–12.

39. Campagna M, Locci E, Piras R, Noto A, Lecca LI, Pilia I, et al. Metabolomic patterns associated to QTc interval in shiftworkers: an explorative analysis. Biomarkers. 2016;21(7):607–13.

40. Lancellotti P, Henri C. The left atrium: an old ‘barometer’ which can reveal great secrets. European journal of heart failure. 2014;16(10):1047–8.

41. Biering-Sørensen T, Kabir M, Waks JW, Thomas J, Post WS, Soliman EZ, et al. Global ECG Measures and Cardiac Structure and Function. The ARIC Study (Atherosclerosis Risk in Communities). 2018;11(3).

42. Tromp J, Khan MAF, Klip IT, Meyer S, de Boer RA, Jaarsma T, et al. Biomarker Profiles in Heart Failure Patients With Preserved and Reduced Ejection Fraction. Journal of the American Heart Association. 2017;6(4).

43. Hunter WG, Kelly JP, McGarrah RW, 3rd, Khouri MG, Craig D, Haynes C, et al. Metabolomic Profiling Identifies Novel Circulating Biomarkers of Mitochondrial Dysfunction Differentially Elevated in Heart Failure With Preserved Versus Reduced Ejection Fraction: Evidence for Shared Metabolic Impairments in Clinical Heart Failure. J Am Heart Assoc. 2016;5(8).

44. Nemutlu E, Zhang S, Xu Y-Z, Terzic A, Zhong L, Dzeja PD, et al. Cardiac Resynchronization Therapy Induces Adaptive Metabolic Transitions in the Metabolomic Profile of Heart Failure. Journal of cardiac failure. 2015;21(6):460–9.

45. Zhang Y, Blasco-Colmenares E, Harms AC, London B, Halder I, Singh M, et al. Serum amine-based metabolites and their association with outcomes in primary prevention implantable cardioverter-defibrillator patients. Europace : European pacing, arrhythmias, and cardiac electrophysiology : journal of the working groups on cardiac pacing, arrhythmias, and cardiac cellular electrophysiology of the European Society of Cardiology. 2016;18(9):1383–90.

46. Cheng ML, Wang CH, Shiao MS, Liu MH, Huang YY, Huang CY, et al. Metabolic disturbances identified in plasma are associated with outcomes in patients with heart failure: diagnostic and prognostic value of metabolomics. J Am Coll Cardiol. 2015;65(15):1509–20.

47. Hanninen SA, Darling PB, Sole MJ, Barr A, Keith ME. The Prevalence of Thiamin Deficiency in Hospitalized Patients With Congestive Heart Failure. Journal of the American College of Cardiology. 2006;47(2):354–61.

48. Shimon H, Almog S, Vered Z, Seligmann H, Shefi M, Peleg E, et al. Improved left ventricular function after thiamine supplementation in patients with congestive heart failure receiving long-term furosemide therapy. The American Journal of Medicine. 1995;98(5):485–90.

49. DiNicolantonio JJ, Lavie CJ, Niazi AK, O’Keefe JH, Hu T. Effects of Thiamine on Cardiac Function in Patients With Systolic Heart Failure: Systematic Review and Metaanalysis of Randomized, Double-Blind, Placebo-Controlled Trials. The Ochsner Journal. 2013;13(4):495–9.

50. Gao K, Zhao H, Gao J, Wen B, Jia C, Wang Z, et al. Mechanism of Chinese Medicine Herbs Effects on Chronic Heart Failure Based on Metabolic Profiling. Frontiers in Pharmacology. 2017;8(864).

51. Wang J, Guo S, Gao K, Shi Q, Fu B, Chen C, et al. Plasma metabolomics combined with personalized diagnosis guided by Chinese medicine reveals subtypes of chronic heart failure. Journal of Traditional Chinese Medical Sciences. 2015;2(2):80–90.

52. Chen J, Li B, Zhao H, Li Z, Wang J, Deng D, et al. Evaluation of Chinese medicine on heart failure based on NMR metabolomics. Journal of Traditional Chinese Medical Sciences. 2016;3(2):100–9.

53. Delles C, Rankin NJ, Boachie C, McConnachie A, Ford I, Kangas A, et al. Nuclear magnetic resonance-based metabolomics identifies phenylalanine as a novel predictor of incident heart failure hospitalisation: results from PROSPER and FINRISK 1997. European journal of heart failure. 2017.

54. Shah SJ, Katz DH, Selvaraj S, Burke MA, Yancy CW, Gheorghiade M, et al. Phenomapping for novel classification of heart failure with preserved ejection fraction. Circulation. 2015;131(3):269–79.

55. Verdonschot JAJ, Hazebroek MR, Derks KWJ, Barandiaran Aizpurua A, Merken JJ, Wang P, et al. Titin cardiomyopathy leads to altered mitochondrial energetics, increased fibrosis and long-term life-threatening arrhythmias. European heart journal. 2018.

56. Ware JS, Cook SA. Role of titin in cardiomyopathy: from DNA variants to patient stratification. Nature reviews Cardiology. 2017.

57. Lee G, Twerenbold R, Tanglay Y, Reichlin T, Honegger U, Wagener M, et al. Clinical benefit of high-sensitivity cardiac troponin I in the detection of exercise-induced myocardial ischemia. American heart journal. 2016;173:8–17.

58. Xanthakis V, Larson MG, Wollert KC, Aragam J, Cheng S, Ho J, et al. Association of novel biomarkers of cardiovascular stress with left ventricular hypertrophy and dysfunction: implications for screening. J Am Heart Assoc. 2013;2(6):e000399.

59. Al-Shareef I, Arabi M, Dabbagh O. Cardiac involvement in nonketotic hyperglycinemia. Journal of child neurology. 2011;26(8):970–3.

60. Shah SH, Sun J-L, Stevens RD, Bain JR, Muehlbauer MJ, Pieper KS, et al. Baseline metabolomic profiles predict cardiovascular events in patients at risk for coronary artery disease. American heart journal.163(5):844-50.e1.

61. Shah SH, Bain JR, Muehlbauer MJ, Stevens RD, Crosslin DR, Haynes C, et al. Association of a Peripheral Blood Metabolic Profile with Coronary Artery Disease and Risk of Subsequent Cardiovascular Events. Circulation: Cardiovascular Genetics. 2010.

62. Ganna A, Salihovic S, Sundstrom J, Broeckling CD, Hedman AK, Magnusson PK, et al. Large-scale metabolomic profiling identifies novel biomarkers for incident coronary heart disease. PLoS genetics. 2014;10(12):e1004801.

63. Ussher JR, Elmariah S, Gerszten RE, Dyck JRB. The Emerging Role of Metabolomics in the Diagnosis and Prognosis of Cardiovascular Disease. Journal of the American College of Cardiology. 2016;68(25):2850–70.

64. Lindahl A, Forshed J, Nordstrom A. Overlap in serum metabolic profiles between non-related diseases: Implications for LC-MS metabolomics biomarker discovery. Biochemical and biophysical research communications. 2016;478(3):1472–7.

65. Chamsi-Pasha MA, Sengupta PP, Zoghbi WA. Handheld Echocardiography. Current State and Future Perspectives. 2017;136(22):2178–88.

66. Gaibazzi N, Baldari C, Faggiano P, Albertini L, Faden G, Pigazzani F, et al. Cardiac calcium score on 2D echo: correlations with cardiac and coronary calcium at multi-detector computed tomography. Cardiovascular Ultrasound. 2014;12(1):43.

67. Gaibazzi N, Porter TR, Agricola E, Cioffi G, Mazzone C, Lorenzoni V, et al. Prognostic Value of Echocardiographic Calcium Score in Patients With a Clinical Indication for Stress Echocardiography. JACC: Cardiovascular Imaging. 2015;8(4):389–96.

68. Montgomery DE, Puthumana JJ, Fox JM, Ogunyankin KO. Global longitudinal strain aids the detection of non-obstructive coronary artery disease in the resting echocardiogram. European Heart Journal - Cardiovascular Imaging. 2012;13(7):579–87.

69. Tada H, Melander O, Louie JZ, Catanese JJ, Rowland CM, Devlin JJ, et al. Risk prediction by genetic risk scores for coronary heart disease is independent of self-reported family history. European Heart Journal. 2016;37(6):561–7.

70. Abraham G, Havulinna AS, Bhalala OG, Byars SG, De Livera AM, Yetukuri L, et al. Genomic prediction of coronary heart disease. European Heart Journal. 2016;37(43):3267–78.

71. Inouye M, Abraham G, Nelson CP, Wood AM, Sweeting MJ, Dudbridge F, et al. Genomic risk prediction of coronary artery disease in nearly 500,000 adults: implications for early screening and primary prevention. bioRxiv. 2018.

72. Khera AV, Chaffin M, Aragam K, Emdin CA, Klarin D, Haas M, et al. Genome-wide polygenic score to identify a monogenic risk-equivalent for coronary disease. bioRxiv. 2017.

73. Khuri-Yakub BT, Oralkan Ö. Capacitive micromachined ultrasonic transducers for medical imaging and therapy. Journal of micromechanics and microengineering : structures, devices, and systems. 2011;21(5):054004–14.

74. Merriman B, Rothberg JM. Progress in ion torrent semiconductor chip based sequencing. Electrophoresis. 2012;33(23):3397–417.

75. Deo RC, Zhang J, Hallock LA, Gajjala S, Nelson L, Fan E, et al. An End-to-End Computer Vision Pipeline for Automated Cardiac Function Assessment by Echocardiography. CoRR. 2017;abs/1706.07342.

76. Gladding P, Schlegel T, Walsh H, Dawson L, O’Shaughnessy B, Scott T. Screening Low Risk Patients Referred for Echocardiography with a 5-min Scout and Advanced Electrocardiography. Heart, Lung and Circulation.26:S28.

77. Nielsen JB, Thorolfsdottir RB, Fritsche LG, Zhou W, Skov MW, Graham SE, et al. Genome-wide association study of 1 million people identifies 111 loci for atrial fibrillation. bioRxiv. 2018.

78. Mints Y, Yarmohammadi H, Khurram IM, Hoyt H, Hansford R, Zimmerman SL, et al. Association of common variations on chromosome 4q25 and left atrial volume in patients with atrial fibrillation. Clinical Medicine Insights Cardiology. 2015;9:39–45.

79. Shoemaker MB, Bollmann A, Lubitz SA, Ueberham L, Saini H, Montgomery J, et al. Common genetic variants and response to atrial fibrillation ablation. Circulation Arrhythmia and electrophysiology. 2015;8(2):296–302.

80. Parvez B, Shoemaker MB, Muhammad R, Richardson R, Jiang L, Blair MA, et al. Common genetic polymorphism at 4q25 locus predicts atrial fibrillation recurrence after successful cardioversion. Heart rhythm. 2013;10(6):849–55.

81. Sauder KA, Skulas-Ray AC, Campbell TS, Johnson JA, Kris-Etherton PM, West SG. Effects of omega-3 fatty acid supplementation on heart rate variability at rest and during acute stress in adults with moderate hypertriglyceridemia. Psychosomatic medicine. 2013;75(4):382–9.

82. Koenig J, Jarczok M, Kuhn W, Morsch K, Schäfer A, Hillecke T, et al. Impact of Caffeine on Heart Rate Variability: A Systematic Review 2013.

83. Richardson T, Rozkovec A, Thomas P, Ryder J, Meckes C, Kerr D. Influence of caffeine on heart rate variability in patients with long-standing type 1 diabetes. Diabetes care. 2004;27(5):1127–31.

84. Notarius CF, Floras JS. Caffeine Enhances Heart Rate Variability in Middle-Aged Healthy, But Not Heart Failure Subjects. Journal of Caffeine Research. 2012;2(2):77–82.

85. Gladding PA, Loader S, Smith K, Zarate E, Green S, Villas-Boas S, et al. Multiomics, virtual reality and artificial intelligence in heart failure. Future cardiology. 2021.

86. Pearl J. Theoretical Impediments to Machine Learning With Seven Sparks from the Causal Revolution 2018.

87. Lund LH, Oldgren J, James S. Registry-Based Pragmatic Trials in Heart Failure: Current Experience and Future Directions. Current Heart Failure Reports. 2017;14(2):59–70.

